# Detection of novel influenza viruses through community and healthcare testing: Implications for surveillance efforts in the United States

**DOI:** 10.1101/2024.02.02.24302173

**Authors:** Sinead E. Morris, Matthew Gilmer, Ryan Threlkel, Lynnette Brammer, Alicia P. Budd, A. Danielle Iuliano, Carrie Reed, Matthew Biggerstaff

## Abstract

**Background:** Novel influenza viruses pose a potential pandemic risk and rapid detection of infections in humans is critical to characterizing the virus and facilitating the implementation of public health response measures.

**Methods:** We use a probabilistic framework to estimate the likelihood that novel influenza virus cases would be detected through testing in different community and healthcare settings (urgent care, emergency department, hospital, and intensive care unit (ICU)) while at low frequencies in the United States. Parameters were informed by data on seasonal influenza virus activity and existing testing practices.

**Results:** In a baseline scenario reflecting the presence of 100 novel virus infections with similar severity to seasonal influenza viruses, the median probability of detecting at least one infection per month was highest in urgent care settings (72%) and when community testing was conducted at random among the general population (77%). However, urgent care testing was over 15 times more efficient (estimated as the number of cases detected per 100,000 tests) due to the larger number of tests required for community testing. In scenarios that assumed increased clinical severity of novel virus infection, median detection probabilities increased across all healthcare settings, particularly in hospitals and ICUs (up to 100%) where testing also became more efficient.

**Conclusions:** Our results suggest that novel influenza virus circulation is likely to be detected through existing healthcare surveillance, with the most efficient testing setting impacted by the disease severity profile. These analyses can help inform future testing strategies to maximize the likelihood of novel influenza detection.

## Introduction

Novel influenza viruses are different from the seasonal influenza viruses currently circulating in humans (A/H3N2, A/H1N1, and B/Victoria). Human infections with novel influenza viruses are generally rare and isolated events that occur through exposure to infected animals (such as livestock) during recreational or occupational activities. At the time of writing (10 May 2024), widespread avian influenza A(H5N1) virus outbreaks occurring among wild and commercial birds since January 2022 have been associated with just two detected human cases of H5N1 in the United States: one individual who was exposed to infected poultry and one who was exposed to infected dairy cattle [1, 2]. The H5N1 viruses associated with these outbreaks do not easily bind to receptors in the human upper respiratory tract and the risk to the general public is currently low [1]. However, a novel influenza virus that transmits efficiently between humans could pose a pandemic risk. Rapid detection of human infection with a novel influenza virus is critical to characterizing the virus causing the infection and facilitating a rapid public health response [3].

Testing is particularly important to distinguish novel influenza virus infection from seasonal influenza or other respiratory virus infections with similar symptom profiles [4]. Although active monitoring and testing of individuals with exposure to infected animals can identify new spillover infections [2], such measures are not designed to detect cases in the wider community following sustained human-to-human transmission. Public health surveillance systems must be equipped to detect novel influenza cases through testing in the community or in healthcare settings where infected individuals might seek care.

We use a probabilistic framework to estimate the likelihood of detection of novel influenza virus cases once sustained human-to-human transmission is occurring at low frequencies within the United States (i.e., 1,000 total cases or less). We consider testing of individuals presenting to different healthcare settings with no known previous exposure to infected animals or humans and use information on testing for seasonal influenza viruses to develop assumptions about plausible testing probabilities. Our findings can help inform testing strategies to improve detection of novel influenza virus cases occurring at low frequencies.

### Methods Model

We adapted an existing framework to estimate detection probabilities for a novel influenza virus in the United States [5]. For a given case of novel influenza virus infection, the probability of detection in a particular healthcare setting can be expressed as

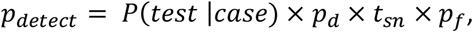

where *P*(*test* |*case*) is the probability that someone is tested in that setting given that they are a case; *p*_*d*_ is the probability that testing occurs while virus is still detectable; *t*_*sn*_ is the test sensitivity; and *p*_*f*_ is the probability a positive test is forwarded to a public health laboratory for further testing. Most commercial assays currently used for human influenza virus testing cannot distinguish novel influenza A viruses from seasonal influenza A viruses. Thus, further testing at a public health laboratory is required for a positive specimen to be identified as a novel virus (until tests specific for that virus become more widely available). We initially assumed 50% of positive specimens are forwarded (i.e., *p*_*f*_ = 50%). This was informed by the average percentage of influenza A hospitalizations that were subtyped between 2010–2019 [6]. However, we considered a range of forwarding levels (25, 50%, 75% and 100%) in sensitivity analyses. All specimens forwarded for further testing were assumed to be correctly identified as a novel influenza virus.

The per case probability of being tested is the combined probability that a case will develop symptoms (*p*_*symp*_), seek care for those symptoms in a particular healthcare setting (*p*_*seek*_), and be tested in that setting (*p*_*test*_), i.e.,

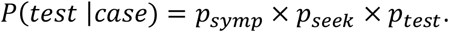

For a certain incidence of novel cases each month, *I*, in a population of size *N* (where *I* is the fraction of the population infected with the novel influenza virus), we estimate the probability of detecting at least one novel case as 1 - the probability of detecting no cases among the entire population, or

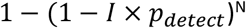

(see the supporting information for further details). The expected number of clinical tests used per month, *E*(*T*), is the combined number of tests conducted among cases and non-cases. Non-cases represent individuals presenting at healthcare settings with respiratory illness symptoms that are not due to novel influenza virus infection. The expected number of tests can be expressed as

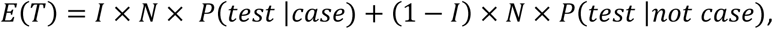

where *P*(*test* |*not case*) is the probability that someone without novel influenza virus infection is tested. The latter quantity is estimated as the background rate of presentation with respiratory illness symptoms to a given healthcare setting among the general population (*b*_*seek*_) multiplied by the probability of being tested in that setting (*p*_*test*_). To compare testing efficiency in different settings we estimated the expected number of detected cases per 100,000 clinical tests conducted as (*I* × *N* × *p*_*detect*_ / *E*(*T*)) × 100,000.

Finally, we considered random testing in the general community as a supplemental strategy that could be deployed in addition to healthcare testing. Given that community testing does not depend on symptom presentation or care-seeking behavior, *P*(*test* |*case*) was simply the frequency of community tests conducted per month and the expected number of tests was *N* × *P*(*test* |*case*). Similarly, *p*_*d*_ was the approximate time (in months) that virus would remain detectable and was parameterized to capture individual variation in virus shedding dynamics. Since community testing would be initiated to seek out novel influenza virus infection, we did not adjust for specimen forwarding (i.e., we assumed all specimens would be tested to distinguish novel influenza virus from seasonal influenza viruses).

For each healthcare and community setting, we drew 10,000 parameter combinations from data-informed distributions (outlined below) and calculated the quantities described above. All analyses and visualizations were generated in R version 4.0.3 using the `data.table`, `truncnorm`, `here`, `scales`, `patchwork`, `colorspace` and `tidyverse` packages [7-14].

### Healthcare settings and model parameterization

We considered three distinct healthcare settings to reflect different care-seeking behaviors and testing practices: (i) outpatient urgent care and emergency departments (UC/ED); (ii) inpatient hospital settings; and (iii) intensive care units (ICU). Each setting was assumed independent such that a person presenting to both (for example, a hospital admission followed by a subsequent ICU admission) could be tested in both, according to the corresponding testing probabilities. Data were collated from various existing influenza surveillance platforms to inform parameters for each setting (Table 1). We defined N = 330 million to approximate the U.S. population [15] and considered incidence values that corresponded to 100 and 1,000 total novel influenza cases.

**Table 1.** Baseline care-seeking and testing parameters. Surveillance platforms are Flu Near You (FNY), Outbreaks Near Me (ONM), VISION Vaccine Effectiveness Network, FluSurv-Net, and IBM MarketScan® Commercial Claims and Encounters Database (MarketScan) [16-19]. Further details of each platform are provided in the supporting information.

| Parameter | Assumed distribution | Source and available timeframe (if applicable) |
| --- | --- | --- |
| Proportion of novel cases developing symptoms, $p_{symp}$ | Uniform with range: 40–80% | [20] |
| Care-seeking and presentation of novel symptomatic cases at specific sites, $p_{seek}$:<br>UC / ED<br>Hospital<br>ICU | Uniform with range:<br>10–20% of symptomatic cases<br>1–2% of symptomatic cases<br>15–20% of hospitalizations | FNY, ONM: 2018–2023<br>CDC burden estimates: 2010-2021 [21]<br>VISION: 2020-2021; FluSurv-Net: 2022–2023 |
| Testing of individuals with ARI, $p_{test}$:<br>UC / ED<br>Hospital<br>ICU | Truncated normal with mean / SD / range:<br>50% / 10% / 10–90%<br>53% / 10% / 20–95%<br>46% / 10% / 1–95% | VISION: December 2021–May 2022 |
| Community testing as a proportion of the general population, regardless of symptoms | 3–6% of general population per month | Assumption following [22] |
| Tests that occur while virus is detectable, $p_d$<br>Healthcare settings<br>Community settings | Uniform with range:<br>50–85%<br>25–50% | Proportion seeking care $\leq 7$ days after symptom onset [23]<br>Proportion of month that virus is detectable [24] |
| Test sensitivity, $t_{sn}$ | Uniform with range 80–100% | [25] |
| Proportion of positive specimens that are forwarded to a public health laboratory, $p_f$ | 50% | Assumption following [6] |
| Background occurrence in the general population, $b_{seek}$, of:<br>ILI*<br>Hospital ARI admissions<br>ICU ARI admissions | Uniform with range:<br>0.6–6%<br>0.03–0.1%<br>0.02–0.03% | FNY, ONM: 2019, 2022<br>MarketScan: 2015–2021<br>MarketScan: 2015–2021 |

Abbreviations: UC = urgent care; ED = emergency department; ICU = intensive care unit; ILI = influenza-like-illness; ARI = acute respiratory illness; SD = standard deviation.
\*Background ILI occurrence is multiplied by care-seeking probabilities in urgent care or emergency departments ( $p_{seek}$ ) to estimate the rate of presentation to urgent care or emergency departments with influenza symptoms in the general population. We omitted data from 2020 and 2021 due to atypically low levels of respiratory virus circulation.

Our baseline scenario reflected a novel influenza virus with similar severity to seasonal influenza. However, we also considered increased severity scenarios that ranged from severity that was similar to COVID-19, to the severity of recent H5N1 virus infections in humans (Table 2). For these scenarios, we assumed similar or increased probabilities of developing symptoms and seeking care in each healthcare setting, while ensuring that the combined percentages did not exceed 100%. We initially assumed testing probabilities were fixed (Table 1) but explored alternative scenarios with increased testing (*p*_*test*_ mean = 90%) to compare the effect of enhanced surveillance across healthcare settings.

**Table 2.**
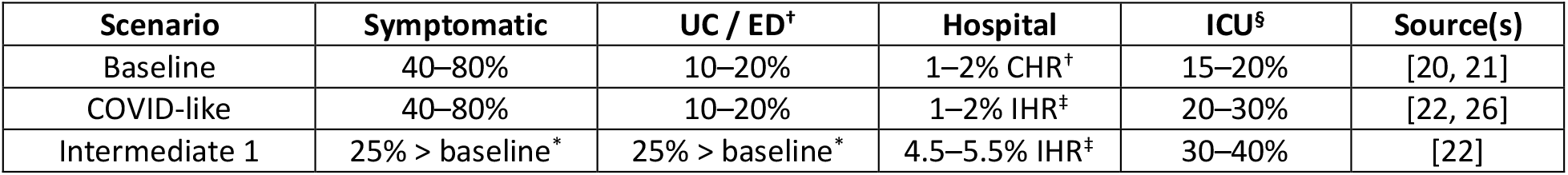
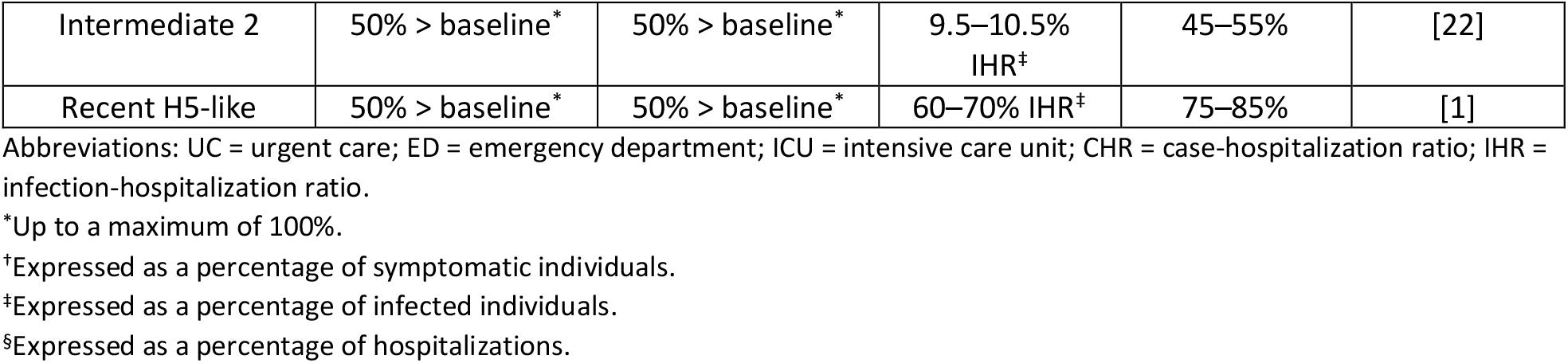
Scenarios for increased symptom severity. All parameters are assumed to follow a Uniform distribution with the reported range.

We also considered scenarios in which testing practices changed according to seasonal influenza activity. For example, clinicians may be less likely to test for influenza viruses during summer months when background respiratory virus activity is low. To explore this, we first defined distinct probability distributions for the background rates of presentation to each healthcare setting, *b*_*seek*_, during peak (November – February) and off-peak (May – August) time periods (Table 3). We then simulated the model for each time period and healthcare setting, assuming the care-seeking behavior of novel influenza cases did not change but that testing in off-peak periods was either equal to, or 50% of, testing in peak periods.

**Table 3.** Baseline occurrence of ILI or ARI symptoms partitioned by peak vs off-peak activity.

| Parameter | Range of uniform distribution | Period | Source and available timeframe |
| --- | --- | --- | --- |
| Occurrence of:<br>ILI* | 1.0–6.0%<br>0.6–2.5% | Peak<br>Off-peak | FNY, ONM: 2019, 2022 |
| Hospital ARI admission | 0.04–0.10% of general population<br>0.03–0.09% of general population | Peak<br>Off-peak | MarketScan: 2015–2021 |
| ICU ARI admission | 0.014–0.035% of general population<br>0.010–0.030% of general population | Peak<br>Off-peak | MarketScan: 2015–2021 |

Abbreviations: ILI = influenza-like-illness; ARI = acute respiratory illness; ICU = intensive care unit; FNY = Flu Near You; ONM = Outbreaks Near Me.
\*Background ILI occurrence is multiplied by care-seeking probabilities in urgent care or emergency departments ( $p_{seek}$ ) to estimate the rate of presentation to urgent care or emergency departments in the general population. We omitted data from 2020 and 2021 due to atypically low levels of respiratory virus circulation.

## Results

We first simulated the model with baseline severity assumptions and no distinction between peak and off-peak time periods. At the lowest incidence (100 novel cases in the population), the median probability of detecting at least one case was highest in community and UC/ED settings, at 77% (95^th^ percentile: 56–91%) and 72% (44–92%), respectively (Figure 1C; Baseline scenario). In comparison, median detection probabilities in hospital and ICU settings were less than 15%. The probability of detection increased across all settings when there were 1,000 assumed novel cases in the population, to 100% (100–100%) in UC/EDs and the community, 74% (47–94%) in hospitals, and 19% (9–35%) in ICUs. Testing in UC/ED settings was always most efficient and detected more cases per 100,000 tests than other settings (Figure 1E). Notably, community testing was least efficient due to the greater number of tests required (more than 10 million per month; Figure 1D), and no setting detected more than 3% of all novel cases under our assumptions (Figure 1F). Increasing the percentage of influenza positive specimens forwarded to public health laboratories to 75% or 100% increased detection probabilities and test efficiency across all healthcare settings (Figure S1). For example, the median detection probability in UC/EDs increased to 85% (58–98%) and 92% (69–99%) at the lowest incidence, respectively. Conversely, a decrease in the percentage forwarded to 25% decreased detection probabilities and test efficiencies, although the relative ordering of setting efficiency was preserved. Thus, for a novel influenza virus with similar severity to seasonal influenza, UC/ED settings are likely to provide greatest opportunities for case detection.

**Figure 1.**
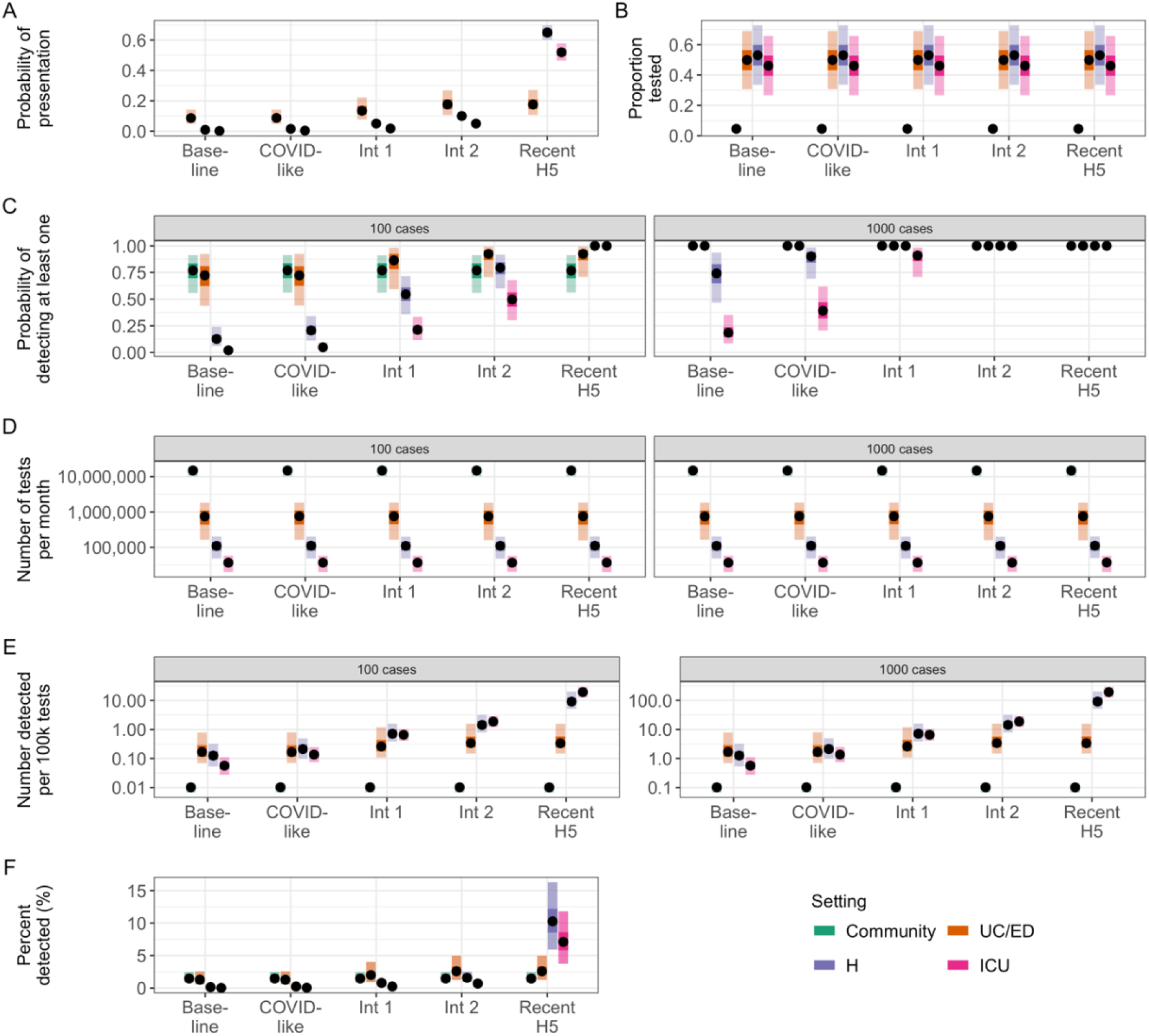
Probabilities of detection and test usage under different severity scenarios. (A) Assumed probabilities of presentation to a particular setting, calculated as *p*_*symp*_ × *p*_*seek*_ for UC/ED, hospital, and ICU settings. All cases are assumed to be in the community, resulting in a probability of one for that setting (not shown). (B) Assumed proportion of individuals with ILI or ARI tested in UC/ED, hospital and ICU settings, or proportion of all individuals tested in the community. (C) Estimated probability of detecting at least one novel case per month. Panels indicate different assumed levels of incidence (100 and 1,000 novel cases). (D) Expected number of clinical tests used per month. (E) Estimated test efficiency, calculated as the number of detected novel cases per 100,000 tests. (F) Percent of all novel cases detected per month. In all panels, points represent median values across 10,000 simulations, inner shaded bands show 50^th^ percentiles, and outer shaded bands show 95th percentiles. Abbreviations: UC = urgent care; ED = emergency department; H = hospital; ICU = intensive care unit; Int 1 = Intermediate 1; Int 2 = Intermediate 2; ILI = influenza-like illness; ARI = acute respiratory illness.

Given uncertainty in the potential severity of a novel influenza virus, we explored additional scenarios in which cases were more likely to develop symptoms and/or present to a particular healthcare setting than the baseline severity scenario (Table 2; Figure 1A). As severity increased, the probability of detection also increased across all healthcare settings due to the greater probability of requiring medical attention (Figure 1C). The difference between detection probabilities in UC/ED compared with hospital and ICU settings also decreased as cases were more likely to be severe and require admission to the latter. For example, median detection probabilities for ICU settings increased from 2% (1–4%) and 19% (9–35%) at baseline with 100 and 1,000 novel cases, respectively, to 100% (98–100%) and 100% (100– 100%) in the “Recent-H5” scenario. There were also substantial increases in testing efficiency in hospital and ICU settings (Figure 1E) and increases in the percent of novel cases detected (for example, from a maximum of 0.3% in hospital settings at baseline to 16% in the Recent H5 scenario; Figure 1F). Test usage is driven primarily by background seasonal influenza virus testing and thus did not change across severity scenarios (Figure 1D). Simulating an increase in clinical testing probabilities (*p*_*test*_ mean = 90%) substantially increased detection probabilities and test usage for all healthcare settings but did not impact the relative performance among settings (Figure S2).

Finally, we assessed how seasonal changes in background activity could impact probabilities of case detection and testing efficiency. Assuming testing practices did not change seasonally led to equal probabilities of detection in peak and off-peak periods, although testing efficiencies were increased in off-peak periods due to the lower number of background tests conducted (Figure S3). Conversely, assuming a 50% reduction in testing across all healthcare settings in off-peak periods (Figure 2B) reduced the corresponding probabilities of detection (Figure 2C). However, for the most severe scenarios (Intermediate 1, Intermediate 2, and Recent H5) there was always at least one healthcare setting with a median detection probability greater than 60% in off-peak periods at the lowest incidence.

**Figure 2.**
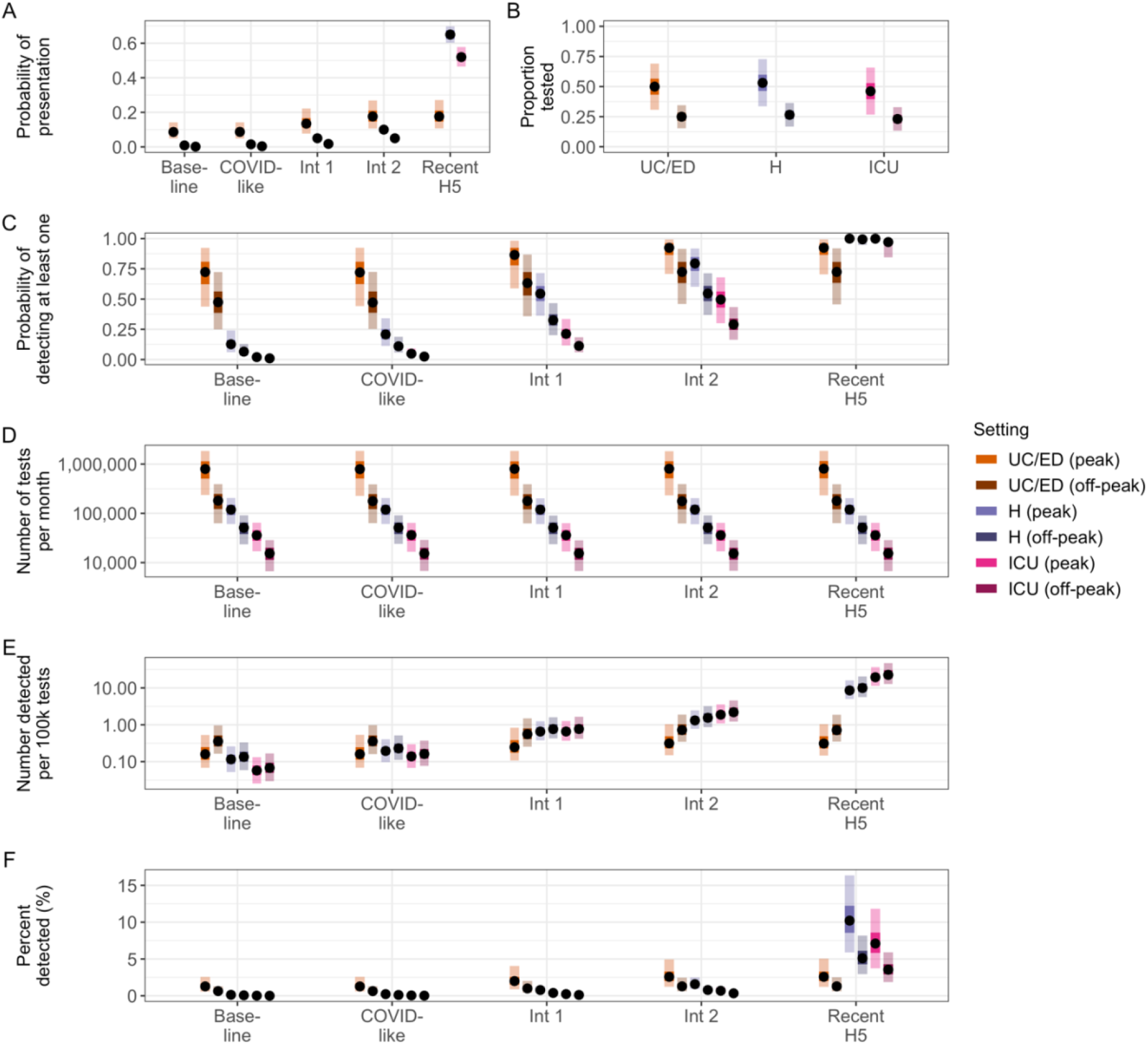
Probabilities of detection and test usage in healthcare settings assuming reduced testing probabilities during periods of off-peak seasonal activity. Incidence is fixed at 100 novel cases in the population. (A) Assumed probabilities of presentation to a particular setting, calculated as *p*_*symp*_ × *p*_*seek*_. Ranges are constant in peak and off-peak periods. (B) Assumed proportion of individuals with ILI or ARI tested in peak and off-peak periods. Ranges are constant across severity scenarios. (C) Estimated probability of detecting at least one novel case per month. (D) Expected number of clinical tests used per month. (E) Estimated test efficiency, calculated as the number of detected novel cases per 100,000 tests. (F) Percent of all novel cases detected per month. In all panels, points represent median values across 10,000 simulations, inner shaded bands are the 50^th^ percentiles, and outer shaded bands are the 95th percentiles. Abbreviations: UC = urgent care; ED = emergency department; H = hospital; ICU = intensive care unit; Int 1 = Intermediate 1; Int 2 = Intermediate 2; ILI = influenza-like illness; ARI = acute respiratory illness.

## Discussion

We modeled the likelihood of detection of novel influenza virus cases occurring at low incidence in the United States. We adapted a simple probabilistic framework that accounted for symptom severity, care-seeking behavior, and testing practices in different healthcare settings, and used care-seeking and testing information from recent influenza seasons to inform model parameters. We found that the most efficient setting for detection depends on the severity profile of the novel influenza virus. Although the percent of total novel influenza cases detected was relatively low, the probabilities of detecting at least one case, and thus identifying novel influenza virus circulation, were high in at least one setting across a range of different testing, severity, and specimen forwarding assumptions.

The high probabilities of detecting at least one case that we have estimated here are relevant for public health pandemic preparedness. The detection of one case would facilitate the implementation of public health actions including increased testing strategies, further virus characterization, vaccine development (if warranted), the implementation of appropriate public health control measures, and updated recommendations for the use of influenza antiviral medications. One key parameter influencing the detection probability was the probability of testing in each healthcare setting. We found that detection probabilities could decrease if influenza testing is substantially reduced below in-season values (for example, during off-peak months). However, it is also possible that clusters of cases and outbreaks could be more likely to be detected and tested during off-peak months if clinicians remain vigilant for evidence of atypical respiratory virus signs or symptoms. The detection probability was also influenced by assumptions about the forwarding of clinical specimens. Our baseline value of 50% was informed by subtyping information from hospitalized influenza infections between 2010–2019 [6]. However, we included a lower bound of 25% to reflect recent post-COVID-19 pandemic trends and potentially reduced forwarding in UC/ED outpatient settings [18]. We also included higher values up to 100% to explore maximum attainable detection probabilities if all tests were forwarded and found a substantial improvement in our estimates. Therefore, during the current H5N1 situation, it is critical that clinicians maintain high testing frequencies and forward influenza A positive specimens to public health laboratories for further testing when recommended. Finally, given the severity of prior H5N1 cases (for example, there has been a 50% case-fatality proportion in cases identified since 1997 [1]), additional strategies to increase testing in ICU settings may help increase the likelihood of detection and testing efficiency, particularly during summer months when background acute respiratory illness rates are low.

Although the probability of detecting one case was generally high, the percent of total cases detected was low, especially in the lower severity scenarios. This finding assumes there are no immediate changes to testing or healthcare seeking behavior once the first case is detected, and arises because detection of influenza through clinical settings requires someone to become symptomatic, seek care, be tested in a timely manner, and have a positive specimen forwarded for further characterization. Although community testing removes these barriers to identification, it is resource intensive and would need to occur even in the absence of perceived novel influenza virus spread to be effective, potentially requiring over 100 million tests per year at the level modeled in this analysis. Similarly, at-home or self-administered tests could alleviate issues associated with care-seeking and clinical testing practices. However, such tests would need to be specific to the novel influenza virus and undergo potentially lengthy development and authorization procedures before being available for widespread use. Pandemic planning efforts should therefore include strategies to rapidly increase testing of acute respiratory illness cases in clinical settings once human-to-human spread of a novel influenza virus has been identified or is likely. Such strategies should account for the possibility that many cases may not be detected, even with increased testing.

There are several caveats to our modeling framework. First, as our primary aim was to estimate detection capabilities once sustained human-to-human transmission is occurring within the United States, we did not consider surveillance for earlier events that might spark such transmission, such as spillover from infected animals or introductions from outside the United States. It is possible that these events would be associated with a greater probability of testing due to relevant exposure histories, and thus have a greater likelihood of being detected compared with our estimates. Second, we did not stratify detection probabilities by age. The severity of seasonal influenza can vary substantially among different age groups [27], and age patterns of severity may differ for a novel influenza virus compared to seasonal influenza viruses due to immunological imprinting and age-related exposures to previous circulating viruses [28, 29]. Age may also impact testing probabilities and healthcare seeking behavior [30, 31], although mean testing probabilities for children <18 years were similar to those of adults ≥18 years in the VISION data used to parameterize *p*_*test*_ (for example, 55 vs. 50% in UC/ED for children and adults, respectively). Including age in the current framework would require additional assumptions regarding the cross-reactivity of the novel influenza virus with seasonal influenza viruses to infer age-specific severity distributions, and thus reduce the generalizability of our results. Third, we did not explicitly incorporate delays in case admission to hospital or ICU that could reduce the window for viable virus detection relative to other settings. These delays are likely on the order of several days and are captured within our conservative range for the proportion of care-seekers who are tested while virus is still detectable [32]. Fourth, we modeled the United States as a single population and did not explicitly consider spatial or other heterogeneities in care-seeking and testing practices. If such data were available, our analysis could be replicated at finer resolution to assess local response and detection capabilities. Fifth, data were not available to fully inform our test forwarding assumptions. Although we considered a range in sensitivity analyses, further information would increase the accuracy of our detection probability estimates. We also assumed perfect sensitivity and specificity for all forwarded tests in line with evaluation of real-time RT-PCR tests for novel H1N1 variant influenza viruses [33]. Although minor reductions in sensitivity should not substantially impact our detection probability estimates, reductions in specificity could lead to false positive results that we have not considered. However, the number of false positive results is likely to be small unless testing reaches extremely high levels, such as considered here in the community setting.

Finally, our assumed inputs for baseline testing and background activity were informed by previous influenza seasons and may not reflect future changes to these values. Where possible, we developed parameter distributions based on data from multiple influenza seasons, before and after the COVID-19 pandemic, to account for broad fluctuations in care-seeking behavior, testing practices, and seasonal influenza dynamics. We also explored scenarios with increased testing to capture the potential impacts of changes to healthcare surveillance following additional policy recommendations. More generally, our estimates of detection probabilities and test efficiency reflect the combined uncertainty in each underlying parameter value and should thus be robust to small changes in any single parameter.

Novel influenza viruses pose a potential pandemic risk, and prompt detection is critical to characterizing the virus causing the infection and facilitating a rapid public health response. Here we demonstrate how a simple probabilistic framework can be used to estimate novel influenza virus detection probabilities through testing in different community and healthcare settings, and can help inform the targeting of future testing efforts. Our work was motivated by the 2022–2024 H5N1 situation in the United States but could be applied more broadly to other locations and/or other potential novel influenza viruses.

## Data Availability

All model inputs and code needed to perform the analysis are available at https://github.com/CDCgov/novel-flu-detection. Data were used solely to inform model inputs and were the result of secondary analyses; the original sources are cited in the text.

## Author Contributions

**Sinead E. Morris:** Conceptualization; Formal analysis; Investigation; Methodology; Visualization; Writing original draft; Writing - review and editing. **Matthew Gilmer**: Data curation; Writing - review and editing. **Ryan Threlkel**: Data curation; Writing - review and editing. **Lynnette Brammer:** Conceptualization; Writing - review and editing. **Alicia P. Budd**: Conceptualization; Writing - review and editing. **A. Danielle Iuliano**: Data curation; Writing - review and editing. **Carrie Reed**: Conceptualization; Investigation; Supervision; Writing - review and editing. **Matthew Biggerstaff**: Conceptualization; Investigation; Methodology; Supervision; Writing - review and editing.

