## Supporting Information for "Detection of novel influenza viruses through community and healthcare testing: Implications for surveillance efforts in the United States"

### Probability of detecting at least one case

We define $p_{detect}$ to be the probability of detection, given that someone is a case of novel influenza virus. If $I$ is the incidence of novel influenza virus in a population of size $N$ (i.e., $I$ is the fraction of the population infected with the novel influenza virus), then $I \times p_{detect}$ is the probability of detection for an average person in the population and $1 - I \times p_{detect}$ is the corresponding probability for no detection. It follows that $(1 - I \times p_{detect})^N$ is the probability that there are no detections among the entire population, and so $1 - (1 - I \times p_{detect})^N$ is the probability that there is at least one detection.

### Surveillance systems

#### Flu Near You (FNY) and Outbreaks Near Me (ONM)

FNY and ONM are participatory surveillance systems in which individuals can self-report a variety of respiratory virus signs and symptoms in addition to care-seeking behavior and testing results. ONM was launched during the 2020/21 influenza season and is the successor to FNY. Seasons run from July to June the following year, and the case definition for influenza-like-illness (ILI) is a participant who self-reported fever, chills, or night sweats, in addition to a cough and/or sore throat. In addition to ONM participants, an additional cross-sectional survey among individuals in the United States aged 18 years and older was conducted in collaboration with Momentive, the parent company for Survey Monkey. Individuals who completed an unrelated survey on Survey Monkey were randomly selected and asked to complete a respiratory virus survey containing the same questions as the ONM platform. Responses were weighted to reflect the U.S. population.
For further details see https://outbreaksnearme.org/us/en-US.

#### VISION Vaccine Effectiveness Network

VISION comprises a network of nine sites in eleven states that collect information on influenza-associated outcomes for people of all ages in a variety of healthcare settings including urgent care, emergency departments, and hospitals. The network's primary focus is to assess seasonal influenza vaccine effectiveness against influenza-related illness, but also provides information on testing practices and intensive care unit (ICU) admission for patients presenting with acute respiratory illness.
For further details see https://www.cdc.gov/flu/vaccines-work/vision-network.html.

### FluSurv-NET

FluSurv-NET captures laboratory-confirmed influenza-associated hospitalizations for all ages through a network of acute care hospitals in 14 states, and represents over 29 million people (roughly 9% of the total U.S. population). Surveillance typically runs from October 1st each year to April 30th the following year. Cases are defined as patients who received a positive laboratory-confirmed influenza test within 14 days prior to, or during, hospitalization. For each case, a range of data are collected including any admission to an ICU.
For further details see https://www.cdc.gov/flu/weekly/influenza-hospitalization-surveillance.htm.

### IBM MarketScan® Commercial Claims and Encounters Database (Marketscan)

Marketscan is produced by IBM and comprises de-identified insurance claims data from approximately 40 million people per year through employer-sponsored healthcare plans across all 50 states in the United States. The database includes diagnostic and procedure codes that can be used to identify admissions due to acute respiratory illness (ARI) in different inpatient settings.
For further details see https://www.ibm.com/downloads/cas/OWZWJ0QO.

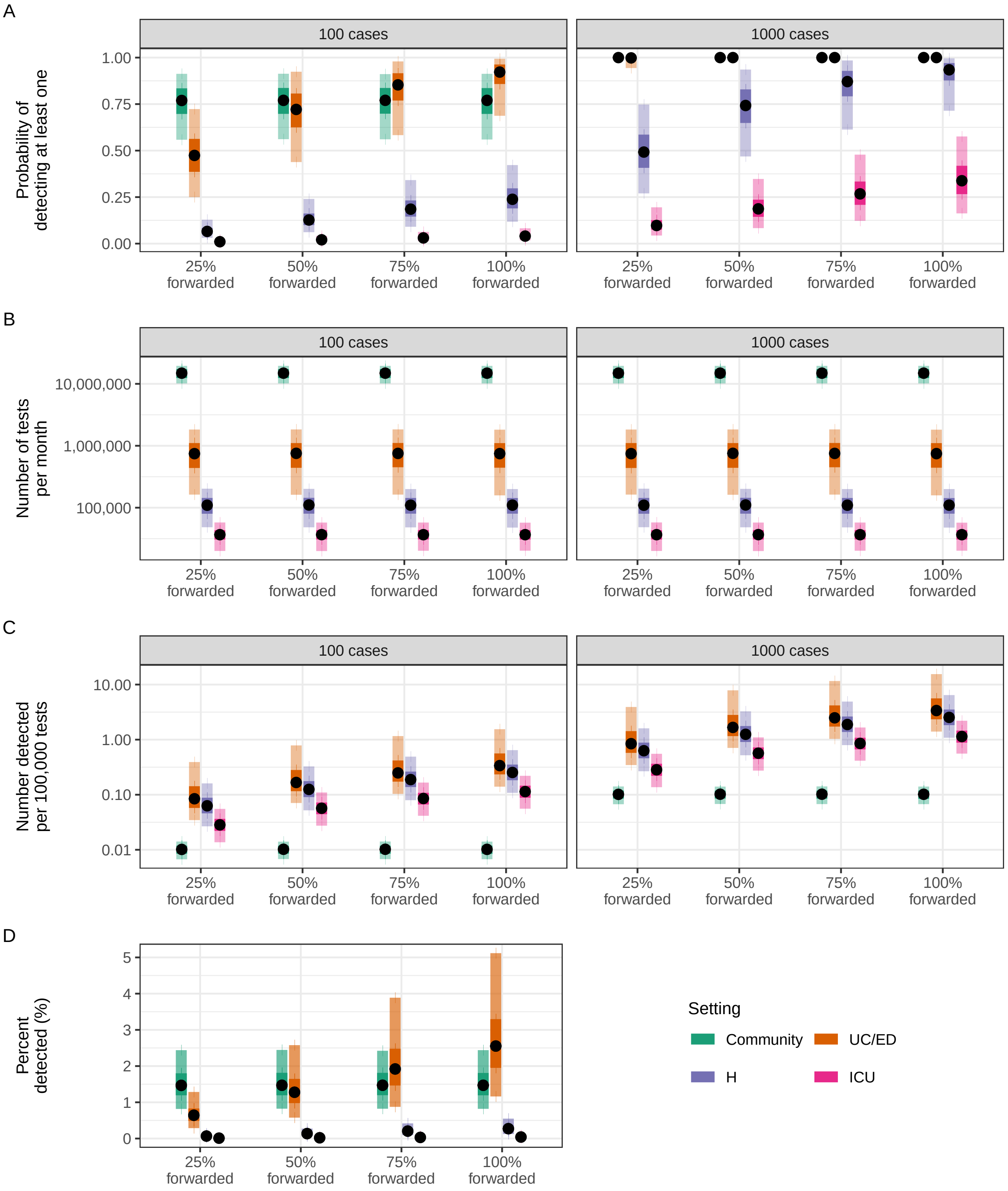


**Figure S1 – Test forwarding, $p_f$, impacts probabilities of detection in healthcare settings under baseline assumptions.** (A) Estimated probability of detecting at least one novel case per month. (B) Expected number of clinical tests used per month. (C) Estimated test efficiency, calculated as the number of detected novel cases per 100,000 tests. (D) Percent of all novel cases detected per month. In all panels, points represent median values across 10,000 simulations, inner shaded bands show 50th percentiles, and outer shaded bands show 95th percentiles. Abbreviations: ED = emergency department; H = hospital; ICU = intensive care unit; UC = urgent care.

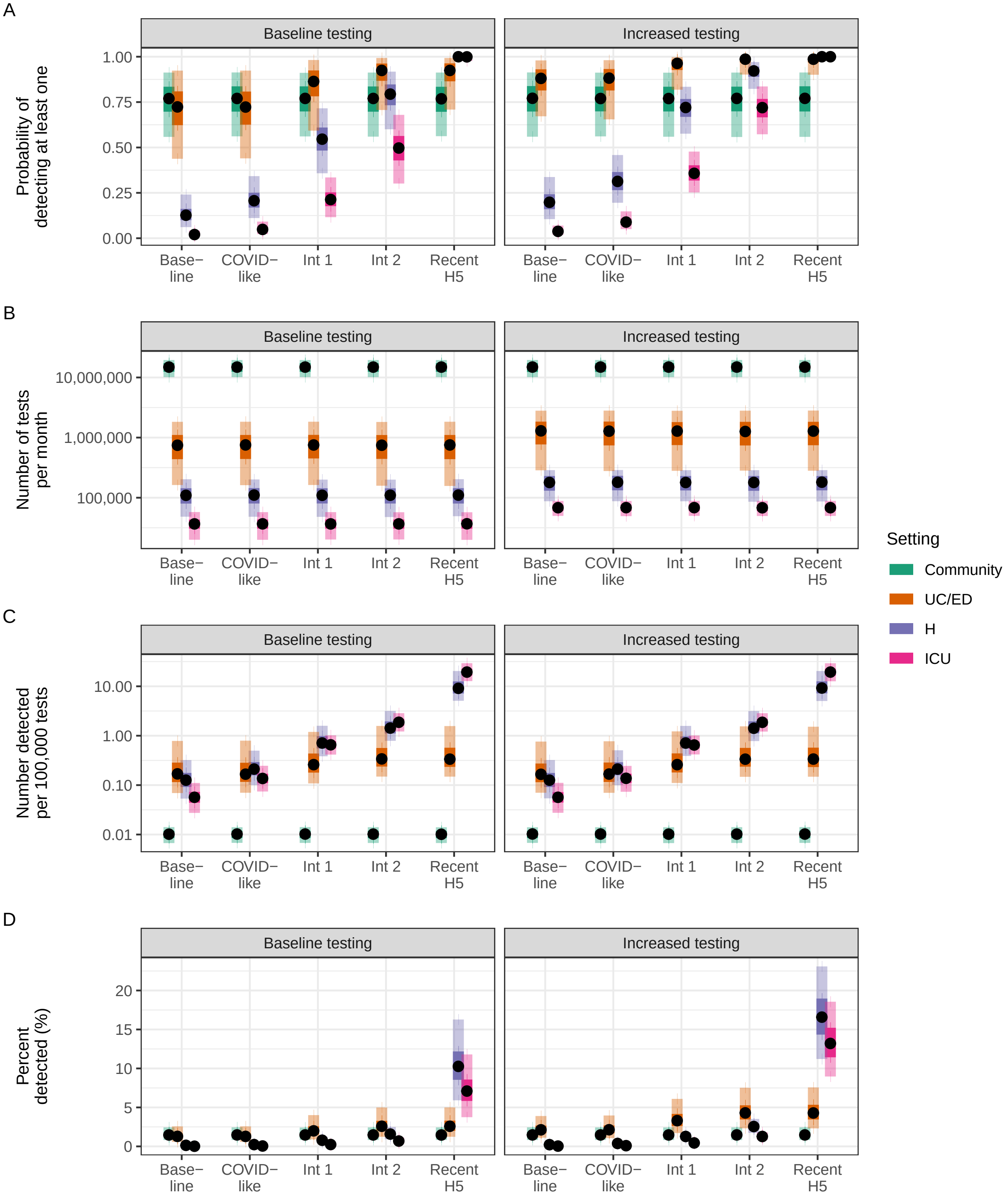


**Figure S2 – Increasing mean healthcare testing probabilities, $p_{test}$, to 90% increases probabilities of detection across all healthcare settings and severity scenarios.** The number of novel cases in the population is fixed at 100. (A) Estimated probability of detecting at least one novel case per month. Panels indicate different assumed testing probabilities (baseline values and increased values). (B) Expected number of clinical tests used per month. (C) Estimated test efficiency, calculated as the number of detected novel cases per 100,000 tests. (D) Percent of all novel cases detected per month. In all panels, points represent median values across 10,000 simulations, inner shaded bands show 50th percentiles, and outer shaded bands show 95th percentiles. Abbreviations: ED = emergency department; H = hospital; ICU = intensive care unit; Int 1 = Intermediate 1; Int 2 = Intermediate 2; UC = urgent care.

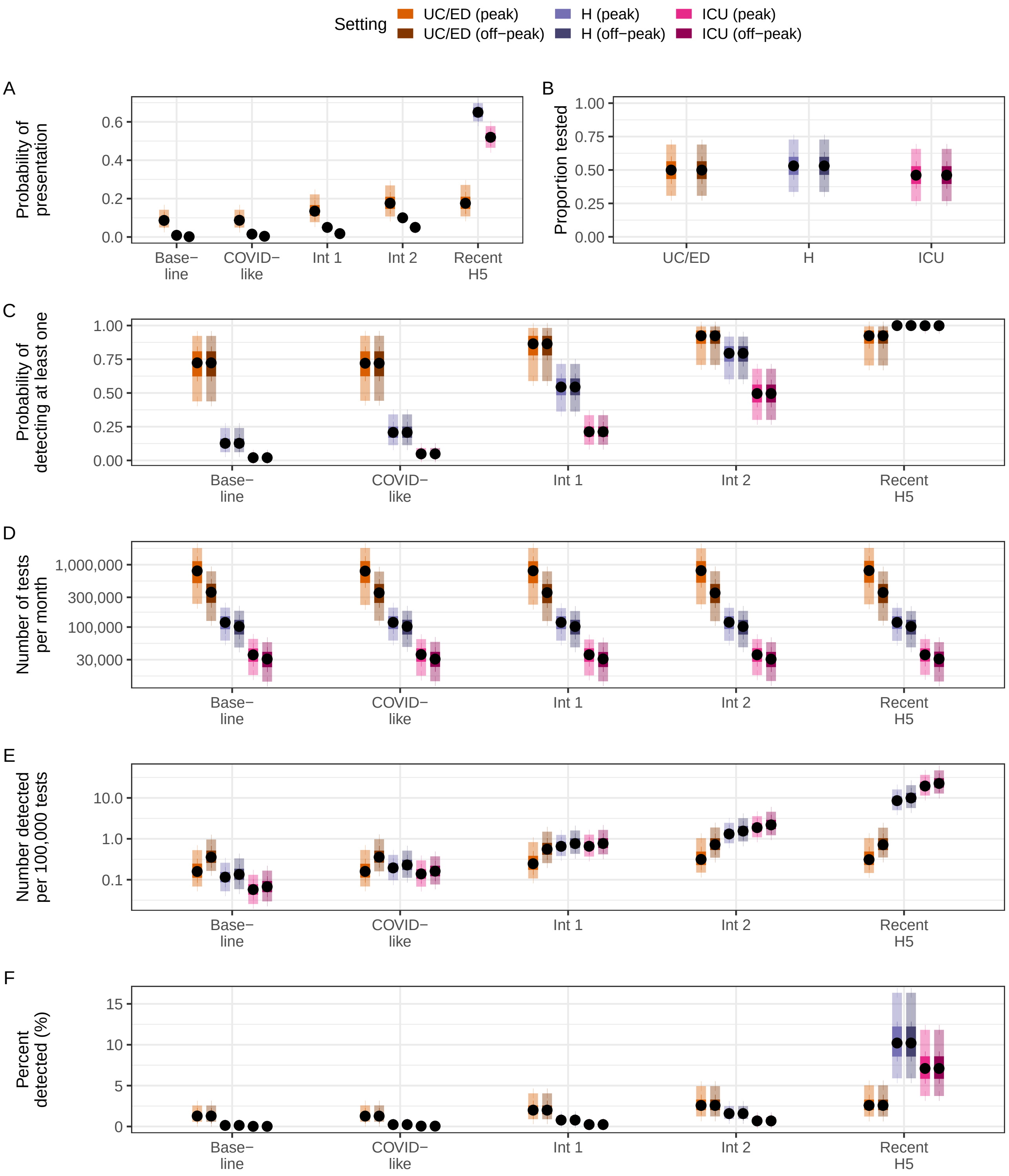


**Figure S3 – Probabilities of detection and test usage in healthcare settings assuming equal testing probabilities in periods of peak and off-peak seasonal activity.** The number of novel cases in the population is fixed at 100. (A) Assumed probabilities of presentation to a particular setting, calculated as $p_{symp} \times p_{seek}$. (B) Assumed proportion of individuals with ILI or ARI tested in peak and off-peak periods. (C) Estimated probability of detecting at least one novel case per month. (D) Expected number of clinical tests used per month. (E) Estimated test efficiency, calculated as the number of detected novel cases per 100,000 tests. (F) Percent of all novel cases detected per month. In all panels, points represent median values across 10,000 simulations, inner shaded bands show 50th percentiles, and outer shaded bands show 95th percentiles. Abbreviations: ARI = acute respiratory illness; ED = emergency department; H = hospital; ICU = intensive care unit; ILI = influenza-like-illness; Int 1 = Intermediate 1; Int 2 = Intermediate 2; UC = urgent care.
